# Prevalence and Predictors of Mechanical Restraint in Involuntary Patients in an Open-Door Inpatient Psychiatric Unit

**DOI:** 10.1101/2025.10.01.25337055

**Authors:** A Fernandez-Ribas, B Jimenez-Fernandez, M Vasquez, L Vargas-Puertolas, E Romero-Diaz, M Delgado-Mari, L Miro-Mezquita, J Cuevas-Esteban

## Abstract

**Purpose:** Mechanical restraint (MR) remains one of the most controversial interventions in psychiatry, particularly in involuntarily admitted patients. This study examined the prevalence and clinical predictors of MR in an open-door psychiatric unit in Spain, a context where national MR rates remain among the highest in Europe.

**Methods:** A retrospective descriptive study of all involuntarily admitted patients (n = 81) was conducted in a 20-bed open-door acute psychiatric unit between June and December 2024. Sociodemographic, diagnostic, and treatment variables were extracted from electronic health records. Bivariate analyses and multivariate logistic regression were used to identify independent predictors of MR.

**Results:** MR was applied in 14.8% of patients, representing a approximately 60% reduction compared with the Spanish national average of 37% reported in multicentre European data. In multivariate analysis, schizophrenia spectrum disorder was the strongest independent predictor (OR = 12.29, 95% CI: 1.75–86.46, p = 0.012). Male gender showed a significant bivariate association but lost significance after adjustment. MR co-occurred with intramuscular psychotropic medication in 91.7% of cases.

**Conclusions:** Even within a high-risk, 100% involuntary population, open-door policies combined with structured de-escalation interventions can substantially reduce MR use. Schizophrenia spectrum disorders emerge as a critical target for prevention strategies. These findings align with convergent evidence from German, Swiss, and Norwegian studies supporting the safety and effectiveness of open-door approaches, and provide the first Spanish data on MR in this model of care.

## Introduction

Mechanical restraint (MR) is one of the most contentious and ethically challenging interventions in modern psychiatry, particularly when applied to involuntarily admitted patients [1,2]. Defined as the use of physical devices to restrict a person’s movement, MR is intended as a last-resort safety measure during acute behavioral crises [1,2]. However, it is associated with substantial risks, including physical injury, psychological trauma, erosion of therapeutic relationships, and violation of human rights principles [1–4]. For involuntary patients—already experiencing loss of liberty—MR represents a “double coercion” with profound ethical implications [3,4].

International evidence shows wide variability in MR prevalence, ranging from 0% to over 50%, influenced by institutional policies, patient populations, and cultural factors [5,6]. In Europe, the multicentre study by Raboch et al. (2010) reported a 37% MR prevalence in Spain—the second highest rate among ten countries [7].

In the Spanish context, several studies provide essential benchmarks. Pérez-Revuelta et al. (2021) conducted an eight-year retrospective analysis in a closed-door acute psychiatric unit, reporting a 12.4% prevalence of mechanical restraint and identifying consistent predictors, including male gender, psychotic disorders, and younger age [8]. More recently, Cañabate Ros et al. (2024) reported strong associations between mechanical restraint and involuntary admission (OR = 17.90), previous restraint episodes (OR = 27.69), and the presence of positive psychotic symptoms (OR = 1.35) [9]. In our own healthcare region, El-Abidi et al. (2021) found a 25% MR prevalence in a mixed voluntary/involuntary acute unit, further emphasizing the need for reduction strategies in routine clinical practice [10].

Open-door psychiatric units, increasingly implemented across Europe, aim to enhance autonomy and recovery by removing locked doors and allowing supervised mobility [11,12]. Large-scale studies—such as Schneeberger et al. (2017) in Germany—have shown significant reductions in seclusion and restraint in open-door hospitals compared to locked wards (OR = 0.369, p = 0.002) without increases in aggressive incidents [13]. More recently, Indregard et al. (2024) conducted the first randomized controlled trial in Norway, reporting lower coercion prevalence in open-door wards (26.5%) versus treatment-as-usual (33.4%), and MR rates of 3.3% versus 4.8%, respectively [14].

Despite the WHO’s QualityRights initiative advocating for the elimination of coercion and the global shift toward rights-based mental health care [15], few studies have examined MR prevalence and predictors specifically in open-door settings with exclusively involuntary populations.

This study aims to determine the prevalence and identify independent predictors of MR in an exclusively involuntary cohort admitted to an open-door psychiatric unit in Spain. The findings may inform targeted interventions to reduce coercion while maintaining safety, aligning clinical practice with human rights-based mental health care.

## Methods

### Study Design and Setting

This retrospective descriptive study was conducted between June and December 2024 in the acute open-door psychiatric unit of Germans Trias University Hospital (GTUH), a general multispecialty teaching hospital affiliated with the Autonomous University of Barcelona. Located in Badalona (Barcelona, Spain), GTUH serves a catchment area of 350,530 adults across four community mental health zones within the North Barcelona health district.

At the time of the study, the psychiatric unit operated as a single 20-bed ward, staffed by a multidisciplinary team including psychiatrists, psychiatry residents, nurses, psychologists, social workers, occupational therapists, and auxiliary personnel. The unit provides intensive treatment for severe mental disorders, integrating pharmacotherapy, psychotherapy, occupational therapy, and social interventions. Since 2019, the Safewards care model—comprising ten structured interventions aimed at reducing conflict and containment—has been progressively implemented throughout the unit. Electroconvulsive therapy was available when clinically indicated.

The unit follows a comprehensive open-door policy, allowing patients up to six hours of supervised daily leave from the ward. Approximately 90–95% of admissions occur through the emergency department, with most patients presenting with acute psychiatric symptoms and/or significant risk of self-harm or harm to others. Roughly 40% of admissions are involuntary (court-mandated per Spanish mental health legislation), while 60% are voluntary.

### Sample and Data Collection

The study population comprised all patients admitted involuntarily to the psychiatric unit during the six-month study period. A total of 81 patients met the inclusion criteria. Data were collected retrospectively from electronic medical records and hospital administrative databases, ensuring comprehensive capture of relevant clinical information.

Variables collected included sociodemographic characteristics (age, gender, living arrangements), clinical diagnoses according to DSM-5 criteria, and treatment-related factors (use of intramuscular medication, application of mechanical restraint, substance use at admission and discharge, and history of psychiatric admissions). All diagnostic assignments were made by qualified psychiatrists and verified through chart review.

### Variables and Definitions

The primary outcome variable was the use of mechanical restraint during hospitalization. Mechanical restraint was operationally defined as any use of physical devices to restrict a patient’s movement, documented in clinical records according to institutional protocols. This included restraint episodes initiated prior to ward admission (e.g., in the emergency department), provided they were documented in the medical record.

Independent variables included gender, age, living situation, primary psychiatric diagnosis, history of substance use disorders, administration of intramuscular psychotropic medication, and whether the admission was the patient’s first psychiatric hospitalization. Diagnoses were initially grouped into psychotic disorders, affective disorders, and other disorders. Given prior evidence of their strong association with coercive measures, schizophrenia spectrum disorders were analyzed separately.

### Data Analysis

This manuscript involved the use of a Large Language Model (ChatGPT, OpenAI) to assist with the formulation of scientific text during the writing process. The AI model was employed to support clarity, consistency, and adherence to formal scientific English, under direct human supervision. All content was critically reviewed, edited, and approved by the authors, who maintain full responsibility for the integrity and accuracy of the final manuscript.

Descriptive statistics were utilized to summarize patient characteristics and calculate the overall prevalence of mechanical restraint. Bivariate analyses were conducted using chi-square tests for categorical variables and independent t-tests for continuous variables to identify factors associated with MR use.

Variables demonstrating significance at the p < 0.10 level in bivariate analysis, along with those of established clinical relevance, were entered into a multivariate logistic regression model to determine independent predictors of MR. The final model included gender, age, and schizophrenia spectrum disorder diagnosis. Odds ratios (ORs) with 95% confidence intervals (CIs) and associated p-values were calculated for each predictor. Statistical significance was set at p < 0.05. Analyses were conducted using SPSS version 28.0.

### Ethical Considerations

The study was approved by the Ethics Committee of Germans Trias i Pujol University Hospital (Code: PI-23-245). All data were anonymized prior to analysis, with unique study identifiers replacing patient information. The study complied with the ethical principles outlined in the Declaration of Helsinki and with Spanish national data protection regulations (Ley Orgánica 3/2018 de Protección de Datos Personales).

## Results

### Demographic and Clinical Characteristics

A total of 81 involuntarily admitted patients were included in the analysis. The sample was nearly gender-balanced (49.4% female), with a mean age of 45.1 years (SD = 18.6; range: 18– 89 years). Mechanical restraint (MR) was applied in 14.8% of cases (*n* = 12).

As shown in Table 1, male sex was significantly associated with MR (83.3% in the restrained group vs. 44.9% in the non-restrained group; *p* = 0.014). Patients diagnosed with schizophrenia showed a markedly higher prevalence of MR (33.3% vs. 2.9%; *p* < 0.001).

**Table 1.** Demographic and Clinical Characteristics by Mechanical Restraint Status.

| Variable | No Restraint<br>(n=69) | Mechanical<br>Restraint (n=12) | Total (n=81) | p-value |
| --- | --- | --- | --- | --- |
| Sex, n (%) |  |  |  | 0.014* |
| Female | 38 (55.1%) | 2 (16.7%) | 40 (49.4%) |  |
| Male | 31 (44.9%) | 10 (83.3%) | 41 (50.6%) |  |
| Age, mean (SD) | 46.6 (18.8) | 37.3 (17.2) | 45.1 (18.6) | 0.115 |
| Living Arrangements, n (%) |  |  |  | 0.524 |
| Family | 40 (58.0%) | 3 (25.0%) | 43 (53.1%) |  |
| Alone | 15 (21.7%) | 4 (33.3%) | 19 (23.5%) |  |
| Partner | 7 (10.1%) | 2 (16.7%) | 9 (11.1%) |  |
| Other | 7 (10.1%) | 3 (25.0%) | 10 (12.3%) |  |
| Substance Use at Admission, n (%) |  |  |  | 0.284 |
| No | 50 (73.5%) | 7 (58.3%) | 57 (71.3%) |  |
| Yes | 18 (26.5%) | 5 (41.7%) | 23 (28.7%) |  |
| First Admission, n (%) |  |  |  | 0.676 |
| No | 41 (60.3%) | 8 (66.7%) | 49 (61.3%) |  |
| Yes | 27 (39.7%) | 4 (33.3%) | 31 (38.8%) |  |

*\*Significant at $p < 0.05$*

No significant differences were found between restrained and non-restrained patients with respect to age (mean 46.6 vs. 37.3 years; *p* = 0.115), living arrangements, substance use at admission, or first psychiatric admission status.

### Diagnostic and Treatment Variables

As displayed in Table 2, broader diagnostic categories (psychotic, affective, or other disorders) were not significantly associated with MR use (*p* = 0.216). However, patients with a diagnosis of schizophrenia had significantly higher odds of being mechanically restrained (*p* < 0.001).

**Table 2.** Diagnostic and Treatment Variables by Mechanical Restraint Status.

| Variable | No Restraint<br>(n=69) | Mechanical<br>Restraint (n=12) | Total (n=81) | p-value |
| --- | --- | --- | --- | --- |
| Primary |  |  |  | 0.206 |
| Diagnosis, n (%) |  |  |  |  |
| Schizophrenia | 2 (2.9%) | 4 (33.3%) | 6 (7.4%) |  |
| Non-specified Psychosis | 20 (29.0%) | 4 (33.3%) | 24 (29.6%) |  |
| Bipolar Disorder | 16 (23.2%) | 2 (16.7%) | 18 (22.2%) |  |
| Depressive Disorder | 10 (14.5%) | 1 (8.3%) | 11 (13.6%) |  |
| Schizoaffective Disorder | 8 (11.6%) | 1 (8.3%) | 9 (11.1%) |  |
| Other Disorders | 13 (18.8%) | 0 (0.0%) | 13 (16.0%) |  |
| Schizophrenia, n (%) |  |  |  | <0.001*** |
| No | 67 (97.1%) | 8 (66.7%) | 75 (92.6%) |  |
| Yes | 2 (2.9%) | 4 (33.3%) | 6 (7.4%) |  |
| Diagnostic Category, n (%) |  |  |  | 0.216 |
| Affective Disorders | 29 (42.0%) | 3 (25.0%) | 32 (39.5%) |  |
| Psychotic Disorders | 34 (49.3%) | 9 (75.0%) | 43 (53.1%) |  |
| Other Disorders | 6 (8.7%) | 0 (0.0%) | 6 (7.4%) |  |
| Intramuscular Medication, n (%) |  |  |  | <0.001*** |
| No | 67 (97.1%) | 1 (8.3%) | 68 (84.0%) |  |
| Yes | 2 (2.9%) | 11 (91.7%) | 13 (16.0%) |  |
| Substance Use Disorder at Discharge, n (%) |  |  |  | 0.107 |
| No | 55 (79.7%) | 7 (58.3%) | 62 (76.5%) |  |
| Yes | 14 (20.3%) | 5 (41.7%) | 19 (23.5%) |  |

\*\*\**Significant at $p < 0.001$*

The use of intramuscular psychotropic medication was strongly associated with MR. Among restrained patients, 91.7% received intramuscular medication compared to 2.9% of non-restrained patients (*p* < 0.001).

Substance use disorder at discharge showed no statistically significant association with MR (*p* = 0.107).

### Multivariate Analysis

As shown in Table 3, schizophrenia remained the only statistically significant independent predictor of MR use after adjustment for age and gender (OR = 12.29; 95% CI: 1.75–86.46; *p* = 0.012).

**Table 3.** Logistic Regression Predicting Mechanical Restraint Use.

| Variable | B | SE | Wald | df | p-value | OR (Exp B) | 95% CI for OR |
| --- | --- | --- | --- | --- | --- | --- | --- |
| Male sex | -1.417 | 0.860 | 2.715 | 1 | 0.099 | 0.242 | 0.045 – |
|  |  |  |  |  |  |  | 1.308 |
| Age | -0.023 | 0.023 | 1.023 | 1 | 0.312 | 0.977 | 0.933 –<br>1.022 |
| Schizophrenia | 2.509 | 0.995 | 6.354 | 1 | 0.012* | 12.291 | 1.747 –<br>86.461 |
| Constant | -0.622 | 0.957 | 0.422 | 1 | 0.516 | 0.537 | — |

*\*Significant at $p < 0.05$*

Although male sex showed a significant association in bivariate analysis, it did not retain statistical significance in the multivariate model (OR = 0.24; 95% CI: 0.045–1.31; *p* = 0.099). Age was not independently associated with MR (OR = 0.98; 95% CI: 0.93–1.02; *p* = 0.312).

## Discussion

This study identified a 14.8% prevalence of mechanical restraint (MR) among involuntarily admitted patients in an open-door acute psychiatric unit. This finding is particularly relevant given that involuntary admission is one of the strongest predictors of MR use across psychiatric settings. By focusing exclusively on this high-risk subgroup, the study provides a stringent test of restraint-reduction strategies within the Spanish healthcare system.

Compared to the 37% national average reported for Spain in the European study by Raboch et al. [7], our observed rate represents a 60% reduction. It is also substantially lower than the 54.9% prevalence reported by Cañabate Ros et al. in a sample of Spanish acute units [9]. Notably, even when compared to more recent data, such as the 12.4% reported by Perez-Revuelta et al. in a mixed sample including voluntary and involuntary patients [8], our results remain comparable despite the higher baseline risk in our population. These comparisons suggest that open-door policies—especially when combined with structured interventions such as Safewards [16] —may contribute to reducing coercive practices in psychiatric care.

Internationally, our findings align with the growing body of evidence supporting the effectiveness of open-door models. The 10-year longitudinal study by Krückl et al. [17] and the multicenter analysis by Schneeberger et al. [13] both reported significant reductions in the use of seclusion and restraint in hospitals with open-door policies. Indregard et al. [14] also demonstrated that patients treated in open-door wards had lower mechanical restraint rates compared to treatment-as-usual wards in a randomized controlled trial. Although direct comparisons are limited by differences in legal frameworks, population characteristics, and definitions of coercion, the convergence of results across healthcare systems strengthens the argument that architectural and policy-level changes can lead to meaningful reductions in coercion without compromising safety.

Furthermore, these international studies underscore the importance of sustained institutional commitment and long-term implementation. In the study by Schneeberger et al., the benefits of open-door policies were observed over more than a decade and across hundreds of thousands of admissions, highlighting the scalability and sustainability of this approach[13]. Similarly, Krückl et al. [17] demonstrated that patients initially admitted to open wards but later transferred to closed settings had a significantly higher likelihood of being subjected to coercive measures, underlining the potential value of maintaining treatment continuity in open settings.

A key clinical finding of this study is the strong association between schizophrenia diagnosis and MR use (OR = 12.29). This diagnostic group accounted for two-thirds of all restrained patients, despite representing less than 10% of the total sample. This aligns with prior evidence identifying schizophrenia and acute psychosis as primary risk factors for coercive measures [18–21]. The strength of this association underlines the need for diagnosis-specific preventive strategies, including early identification of agitation, specialized de-escalation protocols, and targeted psychosocial support from admission onwards.

Another important observation was the high co-occurrence between MR and intramuscular medication administration (91.7%). This suggests that these interventions are often deployed together as part of crisis management protocols. While potentially necessary in acute situations, this practice raises important questions about the availability and effectiveness of alternative de-escalation techniques. Future quality improvement initiatives should explore whether early intervention and structured verbal de-escalation strategies can reduce the reliance on combined physical and pharmacological containment.

Limitations This study has several limitations. First, its retrospective design limits the ability to capture contextual and dynamic factors influencing clinical decision-making. Second, we did not collect data on episodes of psychomotor agitation that were successfully managed without MR. This restricts our ability to identify protective factors or effective de-escalation practices, and may underestimate the broader clinical management of agitation. Third, the sample was drawn from a single center, potentially limiting generalizability to other psychiatric services, particularly those operating under different legal or organizational conditions. Finally, the small sample size may have limited statistical power to detect other relevant predictors.

## Conclusions

Our findings provide novel evidence that mechanical restraint use can be substantially reduced even among high-risk involuntary psychiatric inpatients, when care is delivered in an open-door setting with structured therapeutic interventions. The strong association between schizophrenia and MR highlights the importance of developing targeted preventive strategies for this group. While architectural and policy changes appear necessary, they are not sufficient alone; sustained investment in staff training, structured de-escalation, and diagnosis-specific interventions remains essential. These results support the continued shift toward rights-based, non-coercive psychiatric care in line with international human rights frameworks.

## Data Availability

All data produced in the present study are available upon reasonable request to the authors

## Acknowledgments

The authors thank all participating volunteers for their contribution.

